# Psychological impacts of COVID-19 outbreak in Ethiopia: a systematic review and meta-analysis

**DOI:** 10.1101/2022.07.27.22278107

**Authors:** Tadele lankrew, Belete Gelaw

## Abstract

**Background:** The novel coronavirus disease has led individuals in several medical, psychosocial and economic impacts among the majority of the society such as psychological distress, anxiety, depression, denial, panic, and fear. This pandemic is a disastrous health crisis and becoming a current public health emergency and affects several nations across the world. The widespread of COVID-19 has brought not only the risk of death but also major psychological pressure.

The COVID-19 pandemic led individuals to unavoidable psychological distress, anxiety, depression, denial, panic, and fear. The COVID-19 pandemic is a global public health emergency concern, which is severely affected the community and influences the day-to-day life of individuals in Ethiopia. This systematic review used to investigate the pooled estimate on the psychological impact of COVID-19 in Ethiopia.

**Objective:** The main aim of this systematic review and meta-analysis were to provide comprehensive evidence on the psychological impact of COVID-19 in Ethiopia.

**Methods:** This systematic review and meta-analysis searched through Pub Med, Cochrane Library, Google, Google Scholar, and web of sciences. Data extracted by Microsoft Excel then statistical analyses done using STATA Version 14 software with a random-effects model. The funnel plot checked. The heterogeneity of the studies checked. Subgroup analysis done in relation to the study area and authors’ names.

**Results:** A total of 10 studies with 4,215 participants were included in this systematic review and the overall estimated psychological impact of coronavirus disease in Ethiopia was 42.50% (95% CI (31.18%, 53.81%). According to subgroup analysis, the highest estimated status of the psychological impact of coronavirus disease in Ethiopia are 66.40% and 16.20% in Addis Ababa and Amhara regions respectively.

**Conclusion:** This systematic review revealed that the psychological impact of coronavirus disease in Ethiopia is 42.50%. Multiple education and training and adequate personal protective equipment supplies focusing on the psychological impact of COVID-19 should be avail properly for the community in Ethiopia.

## INTRODUCTION

The novel coronavirus disease (COVID-19) pandemic is the most crucial global health tragedy and the greatest challenge of the humankind that affects the social, economic, and becoming a global health threats(1). Coronavirus Disease-2019 was first detected in December 2019 in Wuhan, China and it has been declared a global pandemic by the World Health Organization (WHO), after it has been rapidly transmitted across the world and results high mortality and morbidity(2).

The outbreak of COVID-19 in Ethiopia was officially recognized on 13 March 2020, after a Japanese national arrived in Ethiopia from his Burkina Faso trip, tested positive for the novel COVID19(3). It is believed that unpredictable negative consequence on psychosocial marked by a sense of uncertainty, confusion and urgency which are serious for the community with its ambiguity of transmission, the intense desire of self-protection, family, and friends, the unknown impact of catching the disease itself, unstoppable spread, the panic and outright misinformation lead to acute stress reaction syndrome(4). It enumerated as a worldwide municipal health emergency due to its hasty spread, an increment of the confirmed case, highly contagious, and high mortality to humans. Following this highly contagious, uncertainties related to the transmission of this outbreak, the intense desire to protect family and friends (and yourself), the irresistible blowout of the pandemic, the restriction of social interaction, and outright misinformation directed the nations across the globe have failed under psychological stress(5).

Health emergency actions like solitary confinement, social distancing, separation from loved ones, the loss of choice, uncertainty concluded causes of the COVID-19 disease create negative psychological impact on the health of the population(6). Attempt of suicide, substantial anger generated and complaints brought following the imposition of quarantine in previous outbreaks(7,8). In the most reviewed studies revealed that quarantine of those suspected with the virus provoke substantial psychological disorder like depression, post-traumatic stress disorder, insomnia, irritability, emotional exhaustion, created serious socioeconomic distress and was found to be a risk factor for both anger and anxiety which remain several months after quarantine(7–9). The high contagious nature of COVID-19 brings, economic loss, loneliness of the infected persons, and fear of the disease leads to psychological problem which became a major concern for global health and leads to inevitable stress, depression, and anxiety that has a significant psychological impact on students, government workers, patients, health workers, and communities around the world(5,10–12). As a result, estimating the pooled psychological impact of the COVID-19 pandemic among the community is important for health authorities to develop preventive strategies and effective treatment modalities to alleviate its negative outcome.

## Methods

The systematic review and meta-analysis were conducted based on the Preferred Reporting Items for Systematic and Meta-analysis (PRISMA) protocols(13) to estimate psychological impact of COVID-19 in Ethiopia. We checked the database (http://www.library.UCSF.edu) and the Cochrane library to ensure this study had not been done before and to avoid duplication. We also checked whether there was any similar ongoing systematic review and meta-analysis in the PROSPERO database. PROSPERO Registration message on April 13, 2021, 2:55 PM; CRD [248395]; reassured that there had been no previous similar studies undertaken.

### Inclusion and exclusion criteria

The inclusion criteria were: (1) participants who are living in Ethiopia, (2) studies that clearly reported the psychological impact of COVID-19 in Ethiopia (3) studies that were conducted in Ethiopia, (4) cross-sectional observational studies, and (5) both published and unpublished studies at any time. Articles were excluded if they were: studies were done outside of Ethiopia, case reports, RCT, review. An attempt was made to contact the corresponding authors using the email address or phone number as provided in the published articles.

### Searching strategy

This search strategy was used to explore all relevant published and unpublished studies about the psychological impact of COVID-19 in Ethiopia in the following databases; Pub Med, Google Scholar, and Cochrane Library were searched. The following core search terms or phrases were used; psychological impact, COVID-19, and Ethiopia. Search terms were predefined to allow a complete search strategy that included all-important studies. All fields within records and MeSH (Medical Subject Headings) and Boolean operators were used to search in the advanced Pub Med and UCSF library search engine. We reviewed studies that assessed the psychological impact of COVID-19 in Ethiopia based on the inclusion criteria through the defined study participants, study period, study design, outcome(s), and response rate of the study in Ethiopia.

### Study selection criteria

The retrieved articles were exported to the reference manager software, EndNote x8, and removed duplicate studies. Three investigators (TL, BT, and KE) independently screened the title and abstract based on established article selection criteria. The details of studies that met the inclusion criteria were imported into the Joanna Briggs Institute’s System for the Unified Management, Assessment and Review of Information (JBI SUMARI, The Joanna Briggs Institute, Adelaide, Australia) critical appraisal tools to evaluate the quality of all studies(14). Two (TL, and KE) independent authors appraised the quality of the studies by criteria adapted for reporting prevalence data and cross-sectional studies. Studies were considered low risk if a score of seven and above of the quality assessment indicators **(Table 1)**. Any disagreements that arose between the reviewers were resolved through discussion with other reviewers.

**Table 1.** Critical appraisal results of eligible studies in the systematic review and meta-analysis on psychological impact of COVID-19 in Ethiopia, 2021.

| Authors | Q1 | Q2 | Q3 | Q4 | Q5 | Q6 | Q7 | Q8 | Q9 | Total |
| --- | --- | --- | --- | --- | --- | --- | --- | --- | --- | --- |
| Abay W., etal | y | Y | y | N | Y | n | y | y | y | 7 |
| Argaw A., etal | y | Y | y | Y | Y | y | y | y | y | 9 |
| Chalachew kassaw | u | Y | y | Y | Y | y | y | y | y | 8 |
| Enyew G., etal | y | Y | y | Y | N | y | y | y | y | 8 |
| Sisay G., etal | y | Y | y | Y | Y | y | y | y | y | 9 |
| Yimenu Y., etal | y | Y | y | Y | Y | y | y | y | y | 9 |
| Yigrem A., etal | y | Y | y | Y | Y | y | y | y | y | 9 |
| Solomon H., etal | y | N | y | N | Y | y | y | y | y | 7 |

Y = Yes, N = No, U = Unclear; JBI Critical Appraisal Checklist for Studies Reporting Prevalence Data: Q1. Was the sample frame appropriate to address the target population? Q2. Were study participants sampled in an appropriate way? Q3. Was the sample size adequate? Q4. Were the study subjects, and the setting described in detail? Q5. Was the data analysis conducted with sufficient coverage of the identified sample? Q6. Were valid methods used for the identification of the condition? Q7. Was the condition measured in a standard, reliable way for all participants? Q8. Was there appropriate statistical analysis? Q9. Was the response rate adequate, and if not, was the low response rate managed appropriately?

#### Data Extraction

Data were extracted by three authors (TL, and BG) using a standardized data extraction format that was developed according to the 2014 Joanna Briggs Institute Reviewers’ Manual. The tool used to extract includes authors’ name, study year, region, study area, and study design, sample size, SNQ, RR, and the proportion of the psychological impact of COVID-19 in Ethiopia.

Articles that fulfilled the predefined criteria were used as a source of data for the final analysis. The reviewers crosschecked it to ensure consistency. Any discrepancy was solved through discussion with other authors and the procedure was repeated to overcome the difference, which resulted during extracting every single study.

### Data analysis

The data were extracted using Microsoft Excel and STATA V. 14 (Stata Corp, College Station, TX, USA) statistical software was used for all statistical analysis. The pooled prevalence of the psychological impact of COVID-19 in Ethiopia was determined with a random effect model. The Heterogeneity among the included studies was checked with forest plot, I^2^ test, and the p-values. Heterogeneity among the included studies was investigated with subgroup analysis. Publication bias was checked with a funnel plot. Subgroup analysis was done by study area and author’s name. The results were presented in the form of text, tables, and figures. Additionally, a univariate meta-regression model was applied by taking sample size, publication year, and quality score of each primary study to investigate the sources of heterogeneity. Finally, a forest plot figure was used to present the point proportions with their 95% CI of the primary studies.

## Results

The combined search strategy identified 981 articles. Of these articles, 22 were excluded due to duplication and 151 articles were fully accessed and assessed for qualification. Eventually, 10 articles met the eligibility criteria and were included in the final meta-analysis **(Fig.1.)**

**Figure 1.**
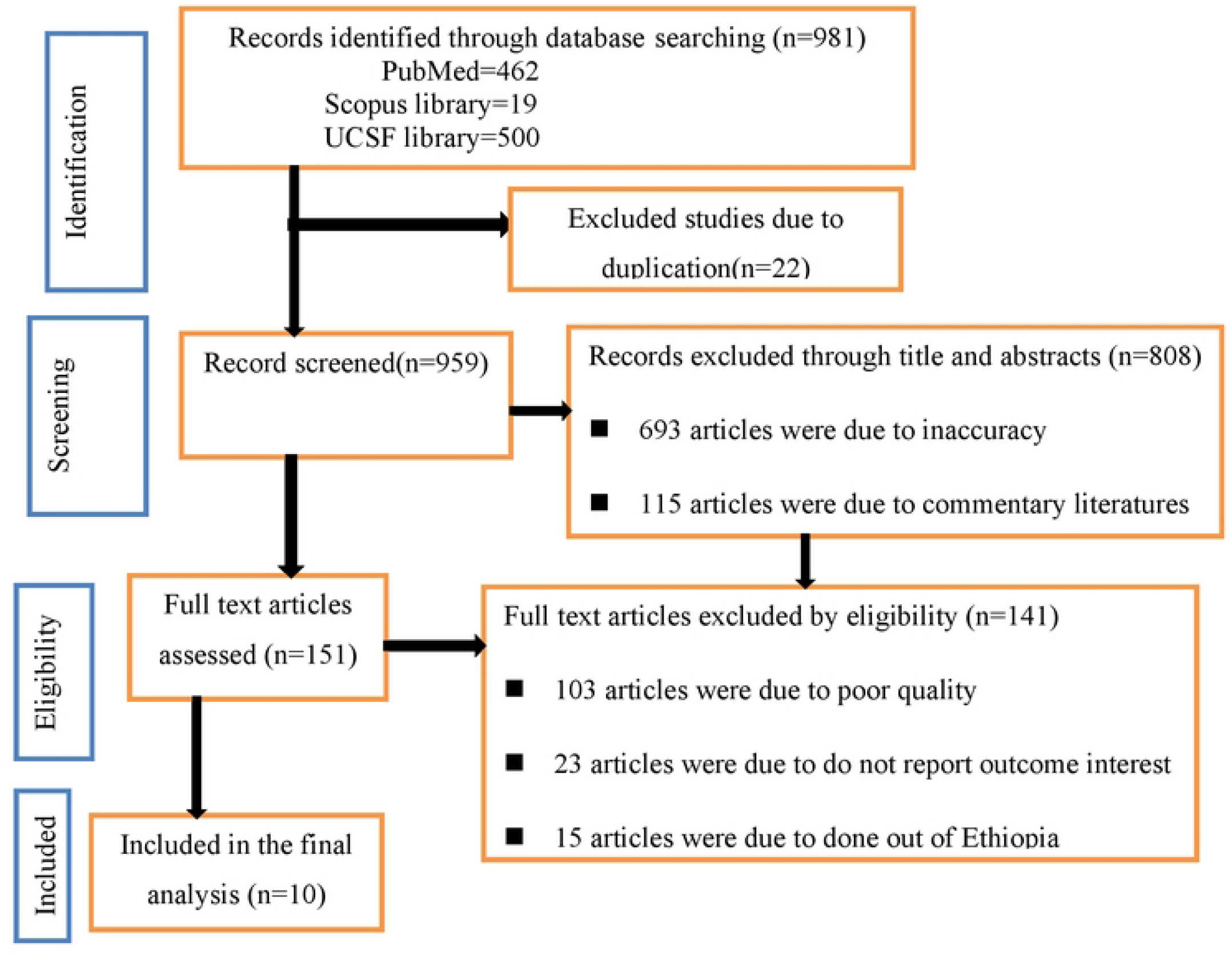
Schematic presentation of study selection for systematic review and meta-analysis for the psychological impacts of COVID-19 preventive measures in Ethiopia, 2021.

### Characteristics of included Studies

This systematic review and meta-analysis include 10 articles with 4,215 study participants. All studies employed a cross-sectional study design. The sample size ranged from 244 to 929, and the response rate ranged from 94% to 100%. The highest and the lowest recoded about the psychological impact of COVID-19 in Ethiopia was (66.6%) and (16.20%) in Addis Ababa and in Amhara region respectively **(Table 2)**.

**Table 2.**
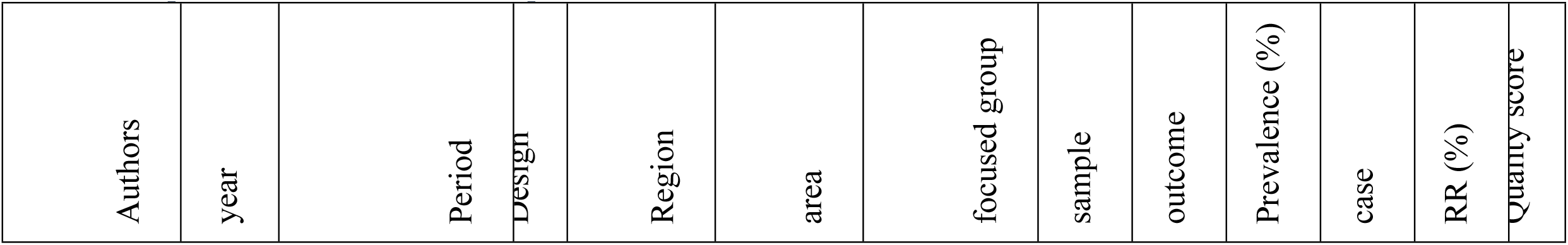
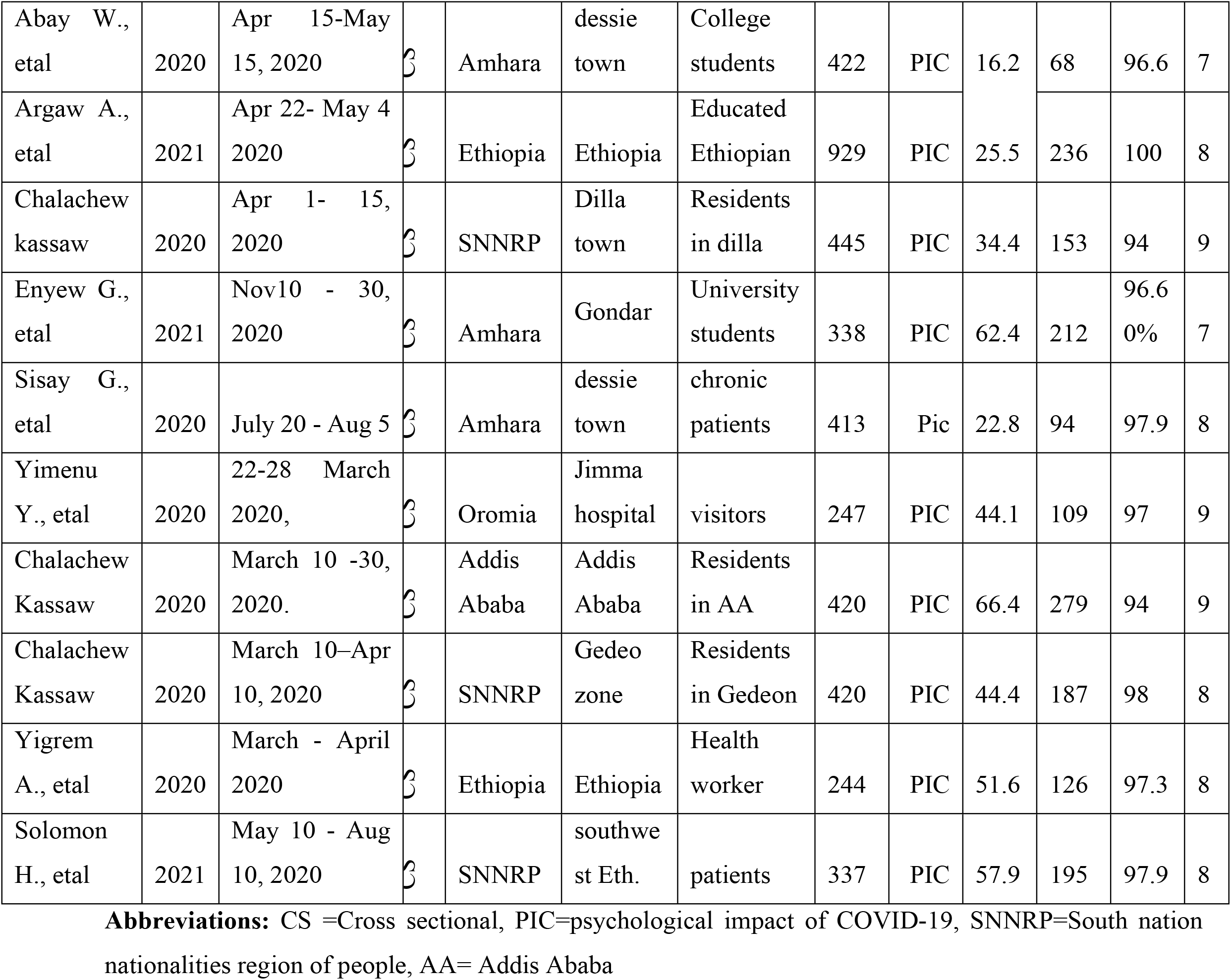
Characteristics of studies included in the systematic review and meta-analysis on psychological impact of COVID-19 in Ethiopia, 2021.

### Publication bias

In this systematic review, the publication bias was checked by funnel plot tests. It gave evidence that the plot resembled symmetric and inverted funnel. Each article’s effective size was allocated against the standard error, and visual inspection of the funnel plot shows that three studies lay on the left side and seven studies on the right side of the line representing the absence of publication bias **(Fig.2)**.

**Figure 1.**
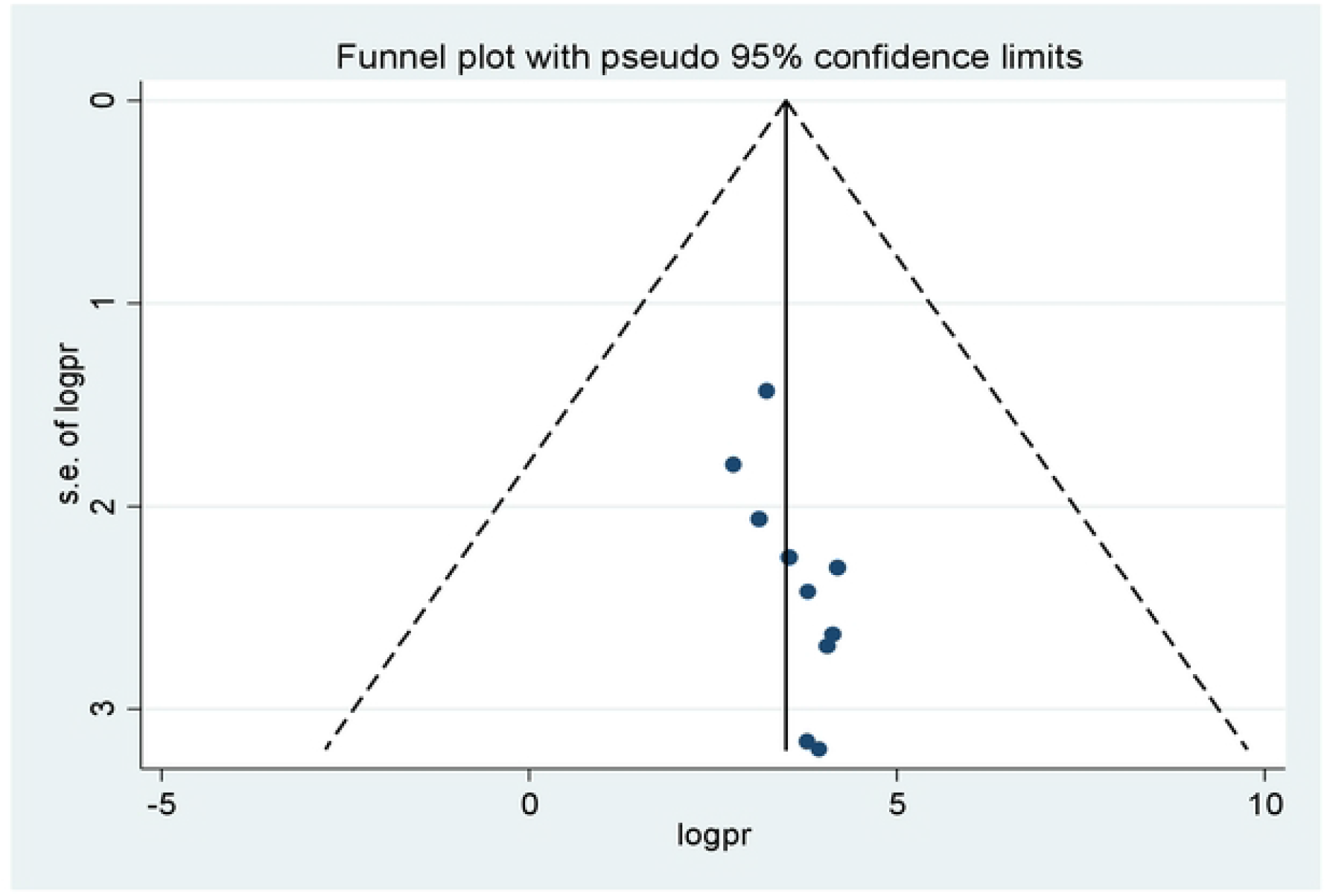
Funnel plots for publication bias of psychological impact of COVID-19 in Ethiopia, 2021.

### Psychological impact of COVID-19 in Ethiopia (a Systematic Review)

By using random-effects model, the estimated status of the psychological impact of COVID-19 in Ethiopia was 42.50% (95% CI (31.18%, 53.81%, I^2^= 98.5%, p ≤0.000) **(Fig.3.)**.

**Figure 1.**
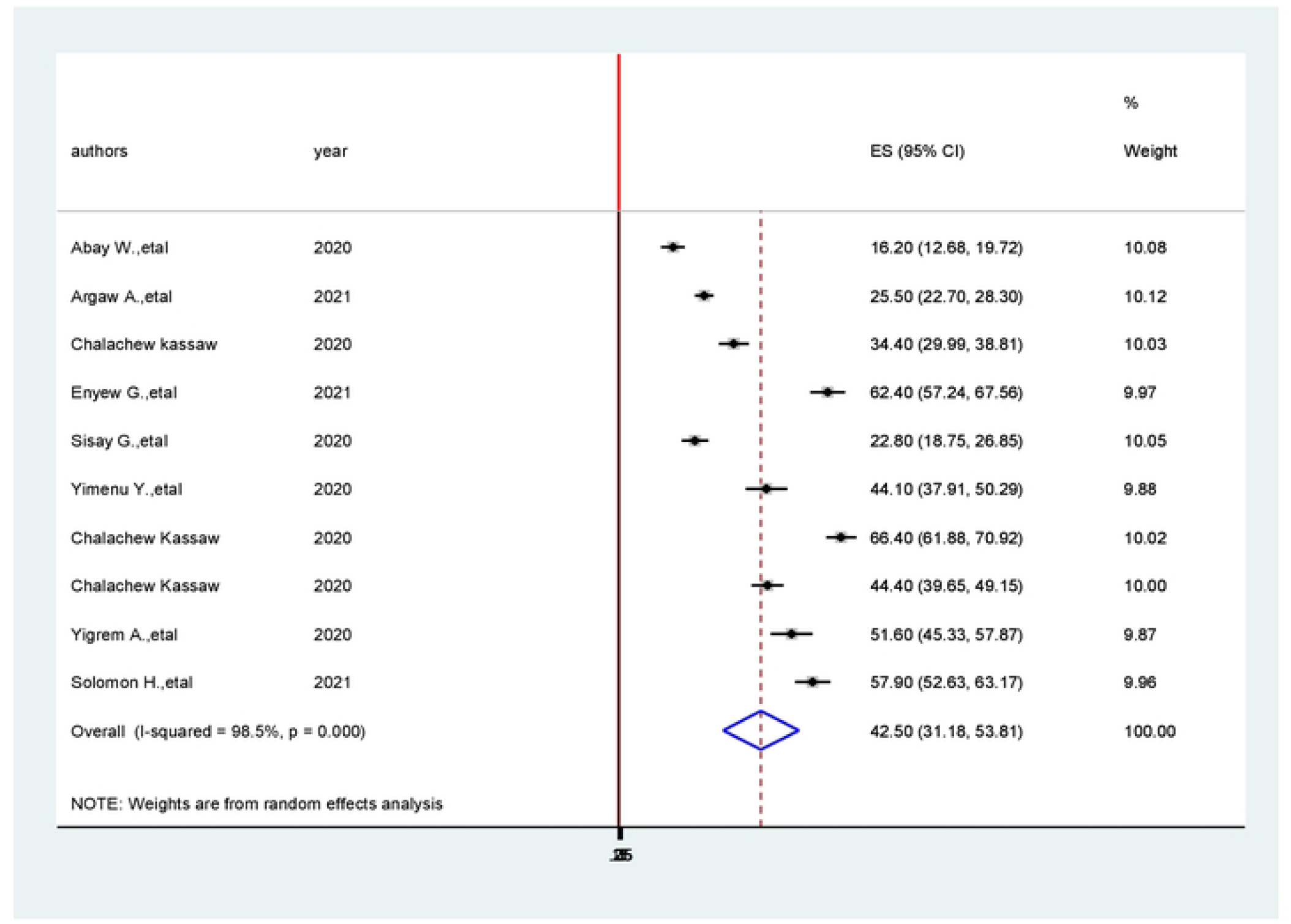
Forest plot for the pooled level of psychological impact of COVID-19 in Ethiopia, 2021.

### Subgroup analysis

Subgroup analysis for the psychological impact of COVID-19 in Ethiopia done based on region and authors’ name. The result revealed that the most and the least prevalent among the 10 primary studies 66.40% (95% CI: 61.88, 70.92%), I^2^= %), and 16.20% (95% CI: 12.68 %, 19.72%), I^2^= 99.10%. p≤0.000) were from Addis Ababa and Amhara regions, respectively **(Fig.4.)**.

**Figure 1.**
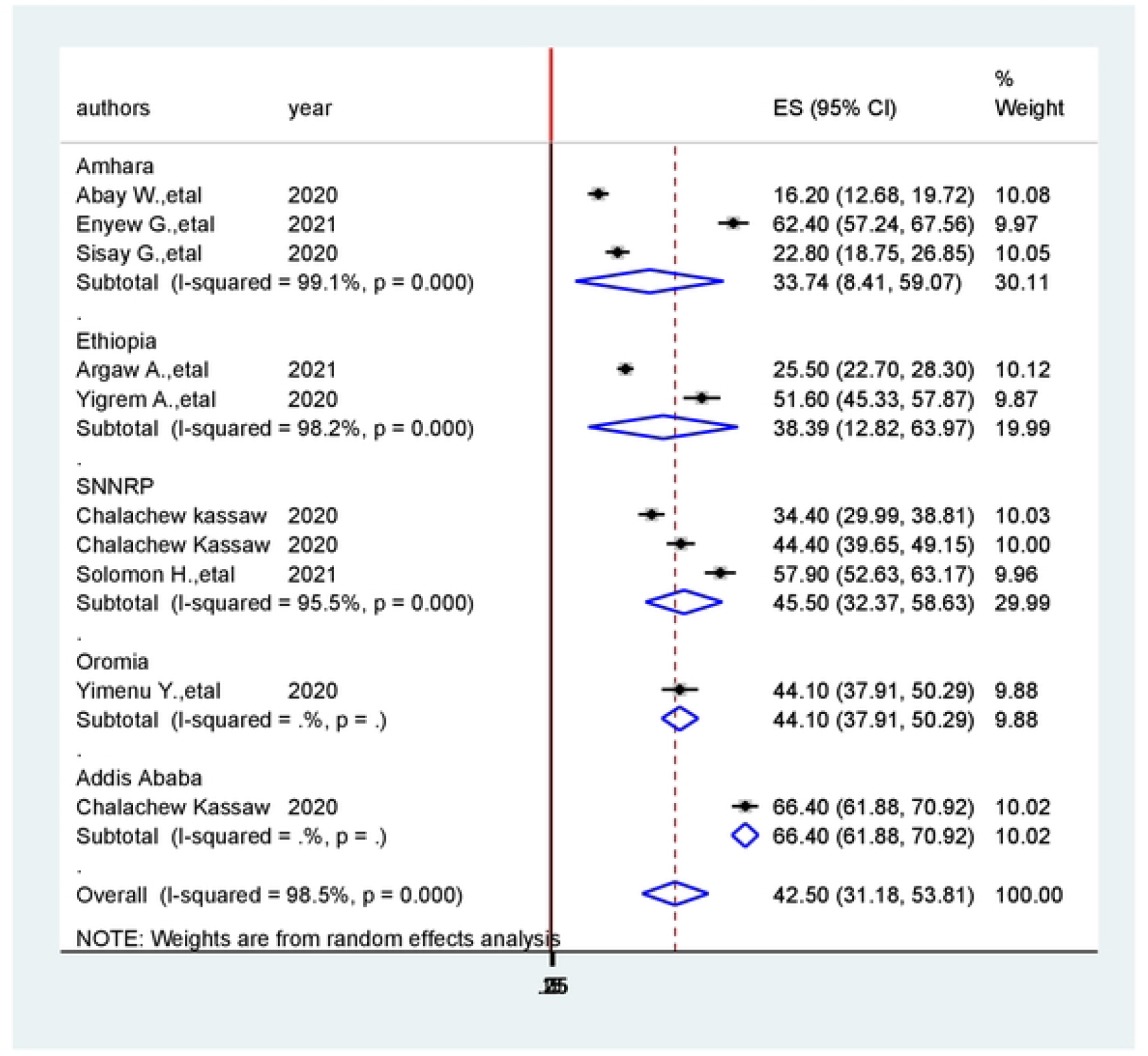
Subgroup analysis on the prevalence of psychological impact of COVID-19 in Ethiopia, 2021

## Discussion

The highly contagious and its widespread transmission of coronavirus, has aggravated the negative psychological impacts among the populations in globally, leading to physical psychological and mental problems in individuals. In the present systematic review and meta-analysis, we found that the COVID-19 pandemic has a significant impact on psychological outcomes. All the articles used in this meta-analysis were cross-sectional studies. Variations in prevalence rates across studies were noticed, which could have resulted from different measurement scales with their different scores. A recent meta-analysis including studies conducted in Ethiopia from the different region of the country reported that the pooled prevalence of psychological impact because of the COVID-19 pandemic in Ethiopia is 42.50%. Sub-group analysis shows that the prevalence of psychological impacts in the population in the Ethiopian context, the result revealed that the most and the least prevalent among the 10 primary studies 66.40% (95% CI: 61.88, 70.92%), I^2^=%), and 16.20% (95% CI: 12.68 %, 19.72%), I^2^= 99.10%; p=0.000) were from Addis Ababa and Amhara regions, respectively.

Using data from 10 cross-sectional studies of 4,215 participants, this current meta-analysis found that the pooled prevalence of psychological impacts in the population in Ethiopia was 42.50% (31.18,53.81). This review is lower than the pooled prevalence of psychological distress/stress which was reported in two studies among the Indian population 43.3% (95% CI: 38.9%– 47.8%) (5). The reason is due the information provided system about Coronavirus was increasing from time onwards and this could be due to the current pandemic among the Indian population having the second-highest number of cases in the world posing a huge challenge on the population who were unprepared. On the other hand the pooled prevalence of psychological stress among the Indian population and the general population in London, UK with in COVID-19 pandemic 34% (95%CI: 20%-50%) (15) and 37.8% (95%CI=30.9–45.5%) (16) were lower than this meta-analysis respectively.

In general, psychological impact of coronavirus directed the community to the making of huge changes to the daily practical activities of the people(17–20). It is a challenge to get adapted to new ways of living such as working from home, temporary loss of income, online schooling of children, and lack of physical contact with other family members, friends and colleagues, and others(21–24). Adapting to this lifestyle changes and concerns related to a disease such as fear of highly contagious of the disease, spreading to near and dear ones, and unable of protecting the vulnerable persons in the family are most of the challenging and stressful task for most people(25–27). This outbreak has triggered social stigma and discriminatory behaviors against people of certain cultural backgrounds like social interaction and those who have been in contact with the virus. Stigma brings serious consequences like stimulating fear, anger, and intolerance directed to other and more likely to experience worse psychological well-being(28–32). The public health response to COVID-19 such as social distancing norms, travel prohibitions, movement restrictions, and quarantines suspected persons, etc. in itself carries the risk of increasing stigma and causing discrimination, which aggravates the psychological impact of coronavirus and affect the population. People in the world are at a higher risk of psychological problems due to multiple factors. These include social isolation; stigma and social discrimination at workplaces and surroundings; higher risk of contracting the disease as they are exposed to patients with high viral load; fear of transmitting the disease to family members; excessive workload with long working hours; inadequate personal protective equipment and high fatality rate of the second-highest number of cases.

The health care professionals are faced with the challenge of assimilating a large amount of information in a short period and acquire new technical skills for properly handling the pandemic. Health authorities must address the concerns of health care professionals and should support them during such major health crises.

In the current systematic review and meta-analysis, we have observed an overall high psychological impact among healthcare workers, the community, and patients with the existing conditions of COVID-19 pandemic. The most common indicators of psychological impact reported across the reviewed studies were stress, anxiety and depression. Common risk factors of heavier psychological burden included being women, being nurses, having high risks of contracting COVID-19, having lower socioeconomic status, social isolation, and spending longer time watching COVID-19 related news; protective factors included having lacking sufficient medical resources, not having up-to-date and accurate health information, and not taking precautionary measures among the people in Ethiopia.

In fact, this meta-analysis was focus comprehensively on the most prevalent psychological problems with which persons have been engaged in Coronavirus epidemics. By collecting and summarizing information about the study subject, the outcomes will provide directions for future researchers and provide information for clinicians for their clinical decision making and health care managers with an understanding of the most prevalent psychological problems in Coronavirus epidemics conditions. More importantly, the findings will give a clear outlook on the priorities of psychological problems in the critical coronavirus epidemic areas specially in Ethiopia. This knowledge will provide rewarding information to inform and support the policymakers in this critical situation. Our results also show that improving family and social support and positive coping strategies are the methods used to reduce risk of psychological distress.

### Strength and limitations

As strength, the study was tried to assess the prevalence of psychological impacts both general populations and health care providers. However, as a limitation, all included studies were cross-sectional, which was difficult to identify causal effect relationships. The other weakness includes the limited number of articles in some regions of Ethiopia, quantitative analysis was not performed and heterogeneity of the articles. Another limitation can be related to the language; we just consider the papers written in the English language.

## Conclusions

In conclusion, this systematic review and meta-analysis revealed that the psychological impact of COVID-19 in Ethiopia was 42.50% (31.18,53.81). Overall, this review suggests that the psychological impacts of Coronaviruses diseases can be widespread significantly in the community. The results of this review indicated that the most prevalent psychological impacts since the first Coronavirus epidemic identified, the community leaders, the healthcare workers and health managers should give priorities for the psychological problems related to COVID-19 in the country.

Health promotions through multiple community instruction, mass media, social media training and by supplying sufficient PPE to the community to prevent the transmission of this pandemic highly recommended to improve the psychological problem of the community. Furthermore, further studies are needed to understand the prevalence and determinant factors of the psychological impact of COVID-19 in Ethiopia using meta-analysis.

Our findings highlight the urgent need for offering mental health services and interventions to target high-risk populations to reduce socioeconomic and gender disparities of psychological distress during the COVID-19 pandemic globally.

## Data Availability

no

no

## Abbreviations

COVID-19: corona virus disease 2019
CDC: Centers for Disease Control and Prevention
WHO: World Health Organization
SNNPR: Southern Nation, Nationalities and Peoples’ of Region

## Data Sharing Statement

Data will be available by request to the corresponding author.

## Authors’ Information

TL, KE, KA, LD and BG are lecturers and practitioner nurses at the School of Nursing and midwifery, College of Medicine and Health Science, Wolaita Sodo University, Ethiopia.

BT, SG are lecturers at the department Nursing and Midwifery, College of Medicine and Health Science, Wolkite University.

## Author Contributions

TL and BG, are involved in the design, selection of articles, data extraction, statistical analysis, manuscript editing, and writing, and take responsibility and be accountable for the contents of the article. All the authors read and approved the final draft of the manuscript.

## Funding

There is no funding to report.

## Disclosure

The authors declare that they have no conflicts of interest for this work.

## Notes

### Competing Interest Statement

The authors have declared no competing interest.

### Clinical Trial

no

### Clinical Protocols

no

